# Predicting Post-Stroke Gait and Balance Function with Simple Neuromotor Measures

**DOI:** 10.1101/2025.06.24.25330026

**Authors:** Margaret Lasonde, Thomas E. Augenstein, Danny Shin, Joshua J. Startup, Edward S. Claflin, James K. Richardson, Chandramouli Krishnan

## Abstract

**Background:** Stroke commonly results in permanent damage to central neural circuits. At a physiological level, this damage manifests as neuromotor impairments like reduced muscle strength, altered coordination, and delayed reaction time. At a functional level, this damage results in reduced gait speed, endurance, and balance ability, which leads to long-term disability and loss of independence. However, the interplay between these neuromotor impairments and functional disability is not well understood. An understanding of these relationships is critical to tailoring rehabilitation approaches for post-stroke recovery.

**Methods:** We measured upper extremity neuromotor capacities as well as mobility and balance measures in 20 chronic stroke survivors and 20 age- and sex-matched controls for this cross-sectional, case-control study. The upper extremity neuromotor capacities included grip strength and various reaction time measures (simple reaction time [SRT], reaction accuracy [All Accuracy], and All Accuracy/SRT) derived using the ReacStick—a novel instrumented ruler-drop test that provides greater ecological validity than computer-based measures. The mobility and balance measures included preferred gait speed, 6-minute walking distance, timed up-and-go test, and single leg balance ability. ANOVAs were used to make between-limb and between-group comparisons, and linear and logistic regression analyses were used to evaluate the neuromotor capacities as predictors of mobility and balance.

**Results:** All neuromotor capacities except All Accuracy and all mobility and balance measures were negatively affected by stroke (all *p*<0.02). We observed that grip strength symmetry (the ratio of grip strength in the paretic limb to the nonparetic limb) was the primary predictor of all mobility measures (all *p*≤0.014), and SRT symmetry and paretic All Accuracy/SRT were the primary predictors of balance (all *p*≤0.002).

**Conclusions:** These results serve as foundational evidence for the relationship between neuromotor performance and functional ability following a stroke and may present an accessible clinical tool for safe measurement of post-stroke mobility and balance.

## Introduction

Stroke is a leading cause of long-term disability, with survivors of stroke often experiencing a variety of motor impairments that impact their ability to perform daily activities.^1^ Among the most common consequences of stroke are deficits in gait, balance, and mobility, which significantly increase the risk of falls and reduce overall quality of life.^2–4^ As a result, individuals with stroke often face challenges in maintaining stability and coordinating movement, which impairs their ability to perform everyday functional tasks such as walking, climbing stairs, or avoiding obstacles.

Relationships between upper limb neuromotor performance and mobility/balance in patients with stroke is of clinical interest because their presence allows the evaluation of mobility/balance while seated, which poses minimal fall risk. There is theoretical and clinical research to suggest that grip strength and grip strength asymmetry may predict mobility/balance. For example, prior research shows that the age-related decline in grip strength relates to the integrity and synchronization of central neural systems rather than a significant change in peripheral muscular factors.^5^ Large cross-sectional studies also show associations between brain structural integrity and grip strength.^6^ If these findings extend to people with stroke, then paretic side grip strength and/or grip strength asymmetry may be useful to clinically quantify degree of diminished central nervous system (CNS) function. Additionally, clinical and epidemiologic research in older (non-stroke) adults have demonstrated that grip strength is a predictor of longevity and fitness.^7^ More recent evidence in older adults finds that greater grip strength asymmetry is associated with diminished gait speed and balance.^8^ In people with a history of stroke, both diminished grip strength and reduced walking speed are associated with mortality from all causes.^9^ However, the relationship between grip strength asymmetry and mobility/balance in people with a history of stroke is unknown.

Reaction time (RT), both simple and complex, offers another means by which upper limb responses may reflect central neurologic integrity and, therefore, functional status. Simple reaction time (SRT) tasks typically involve a single response to a stimulus (e.g., pressing a button when a light appears), while complex reaction time tasks may involve decision-making or response inhibition, such as a go/no-go task. Simple reaction time, which evaluates attention and processing speed, has long been one of the neuromotor measures associated with fall risk.^10^ Simple and complex reaction time measures are commonly assessed using computer-based systems, which provide precise timing and control. Findings from these computer-based tests indicate that stroke survivors often show slower reaction times,^11, 12^ particularly in more complex tasks,^13^ which may contribute to their challenges in performing tasks requiring quick, coordinated movements. While computer-based reaction time measures offer valuable insights, they lack ecological validity, as they do not simulate the real-world demands of motor performance that stroke survivors encounter in daily life. For instance, grabbing a handrail to prevent a fall requires more functional, context-specific assessments of reaction time.

A context-specific and clinically feasible tool for determining both simple and complex reaction times is ReacStick, which is an instrumented rod-shaped device that serves to evaluate reaction time via a ruler drop paradigm (Figure 1). The device has been validated in younger and older people^14–16^ and has been shown to produce greater participant engagement than screen-based measures of reaction time, with shorter and less variable outcomes.^17^ ReacStick parameters have been shown to evaluate mainly executive cognitive functions, which relate to and predict mobility/balance.^18^ As such, ReacStick parameters have predicted unipedal stance time,^19^ grip strength,^20^ cautious gait,^21^ response to lateral and sagittal perturbations,^19, 22^ in community-dwelling older and younger populations,^16, 23^ and populations with clinical conditions including diabetic^19^ and cancer-related neuropathy,^24^ as well as cirrhosis.^25^ However, as with grip strength, the relationships between ReacStick SRT and/or Accuracy and mobility/balance in people with stroke are unknown. Therefore, the purpose of this study was to evaluate how stroke affects grip strength, SRT, and reaction accuracy from go/no go task, and to determine their relationships with commonly used clinical measures of mobility/balance in individuals with a history of stroke. We hypothesized that stroke survivors will exhibit diminished grip strength and greater grip strength asymmetry, along with slower RT, greater RT asymmetry, and reduced reaction accuracy in comparison to age- and sex-matched controls. We further hypothesized that these upper limb neuromotor capacities would significantly predict accepted measures of mobility and balance within the entire cohort and individuals with stroke when analyzed separately.

**Figure 1:**
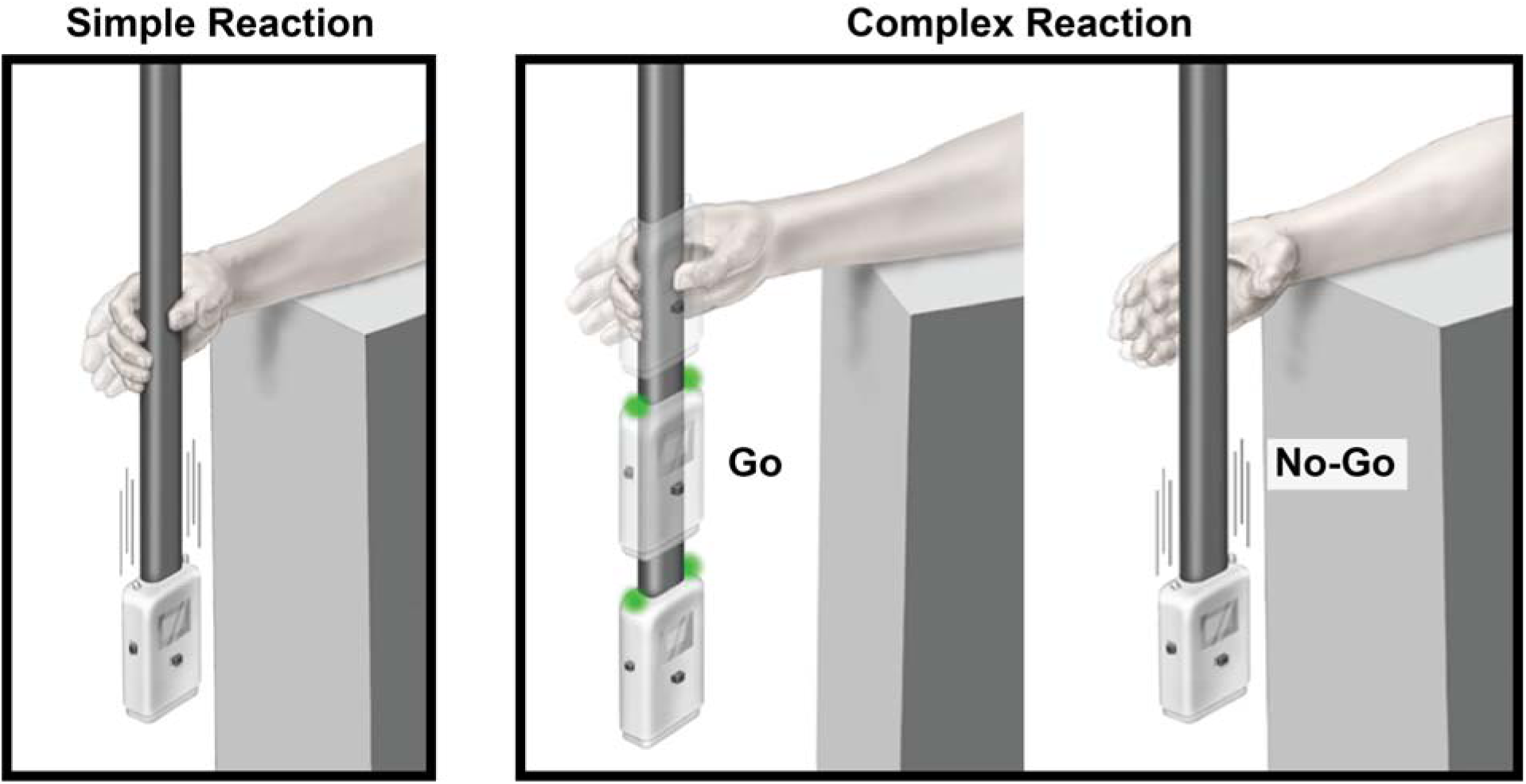
ReacStick is an instrumented rod that can evaluate context-specific reaction time and accuracy, similar to a ruler drop test. When evaluating simple reaction time with the ReacStick (left panel), the examinee waits for the examiner to release the stick and responds by catching the stick as quickly as possible. When evaluating complex reaction time (go/no go, right panel), the examinee only catches the stick when a green “go” light on the stick illuminates

## Methods

### Participants

40 adults (20 individuals with stroke and 20 uninjured controls, Table 1) under the age of 75 years participated in this study. This sample size provided us with a power (1-*β*) > 80% to detect between-group statistical significance with a conservative effect size of d=0.95^26^ at a ≤ 0.05 (G*power 3.1.9.7, Test family: t tests, Means: Difference between two independent means). This sample size also yielded a power greater than >85% to predict mobility measures with an effect size f^2^=0.75^24^ using multiple linear regression analyses (Test family: F tests, Linear multiple Regression: Fixed model, R^2^ deviation from zero, 13 predictors).

**Table 1:**
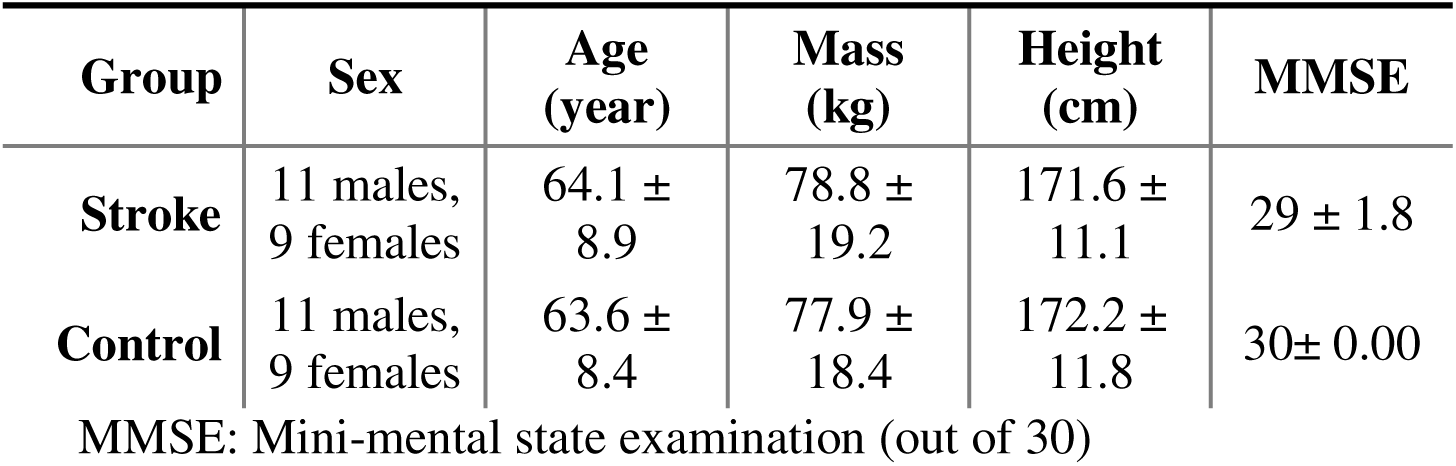
Demographics of participants.

Stroke survivors were included in the study if they (1) had a radiologically (CT or MRI) confirmed ischemic or hemorrhagic stroke at least 6 months prior to the study, (2) had no significant cognitive deficits (Mini-Mental State Examination [MMSE] Score ≥ 22)^27^, (3) had no documented major sensory or proprioceptive deficits, (4) were able to walk independently with or without assistive devices, (5) had no history of uncontrolled diabetes or hypertension, and (6) had no major orthopaedic conditions (e.g., joint replacements) that could interfere with testing. Control participants were included in the study if they (1) had no significant cognitive deficits (MMSE ≥ 22), (2) had no significant orthopaedic or neurological conditions, and (3) had no history of major medical conditions, including uncontrolled diabetes or hypertension. Each stroke survivor was age- and sex-matched to an uninjured control participant. All matches were kept within a range of ±5 years, with an average difference of 2.35 years. All participants were recruited and tested between January 2024 and June 2024. Testing was performed at a single laboratory within the University of Michigan, and participants provided written informed consent prior to participation that was approved by the University of Michigan Medical School Institutional Review Board (IRBMED HUM00073356).

### Method for Assessing Grip Strength

Bilateral grip strength was measured using a hand-held dynamometer (Jamar Hydraulic Hand Dynamometer) with analog outputs. The analog outputs of the dynamometer were interfaced with a custom-built circuit that powered the dynamometer and allowed the analog force output to be extracted via a BNC output. A 16-bit National Instruments (NI) Data Acquisition Board (NI-USB-6218) along with a custom-written program in LabVIEW (version 2015) were used to collect the force data. The raw force outputs in voltage were converted into Newtons using equations obtained from a carefully constructed calibration cycle performed prior to testing. During testing, the participants squeezed the device as forcefully as possible for five seconds, repeating this three times on each hand. The maximum force exerted was recorded for each trial and averaged. Three stroke survivors required assistance to position their grip comfortably on the dynamometer.

### Method for Assessing Simple Reaction Time and Reaction Accuracy

Simple and complex (go/no-go) RT measurements were assessed using an instrumented ReacStick.^16^ The ReacStick is a rigid, lightweight bar with a built-in accelerometer, timing circuit, and light-emitting diode (LED) light (Figure 1). During testing, the participants sat with one arm resting on a standard-height table approximately 90 cm above the ground.^14, 16^ The elbow was flexed to 90-degrees, with the ulnar aspect of the forearm in contact with the table and the hand extending over the edge. The examiner stood in front of the participant, suspending the device vertically. Participants positioned their hand so that the spacer box of the device rested between their thumb and other digits. The device was held from the top and suspended vertically prior to release by the examiner.^28^ After each trial, the reset button was used to clear out prior data and start recording new data. Simple and complex reaction time was assessed bilaterally for both the stroke and control groups.

Guidelines for body positioning and grasping of the device were adjusted for the stroke survivor population depending on their individual motor function and range of motion. If a participant was unable to catch a falling device on their affected side, they were instructed to tap the device in front of them instead. Participants who tapped the device did so on both sides, even if there were no functional limitations on their unaffected side, to ensure consistent assessments between sides.

### Simple Reaction Time

Participants were instructed to watch the spacer box between their fingers, and to catch the device as quickly as possible. The device was released by the examiner after random time delays between 2-5 seconds to minimize participant’s potential anticipation of the release. The subject then grasped the falling apparatus as quickly as possible. Each participant completed four practice trials, followed by ten data acquisition trials.

### Reaction Accuracy

To determine reaction accuracy, the device mode was set to complex which caused the light emitting diodes to illuminate randomly during 50% of the trials at the instant the device was released by the examiner. The linear accelerometer sensed the onset of device movement and instantaneously triggered the illumination of the LED.^28^ Neither the participant nor the examiner knew whether the diodes would illuminate prior to each trial. Participants were instructed to catch the device if the light illuminated and to let the device fall to the ground if not. During this evaluation, the accuracy of the response (*i.e.,* go/no-go) was emphasized rather than the speed of the response. Each participant completed at least six practice trials, followed by at least twenty data acquisition trials. Practice trials continued beyond six trials until participants experienced both a light-on and light-off scenario. The examiner did not implement a delay for this variable. Trials with ambiguous outcomes were repeated..

### Mobility Assessments

The baseline mobility assessment began with a 10 Meter Walk Test (10MWT) overground. Participants walked over a 14-meter walkway (with the intermediate 10-meter marked with tapes on the floor) at their preferred walking speed while the examiner recorded timed using a hand-held stopwatch, beginning when the participant’s front foot crossed the start point and stopping when the front foot crossed the end point. Participants completed one practice trial followed by three recorded trials.^29^

Following the 10MWT, participants completed a six-minute walk (6MWT) test. Participants walked at a safe and comfortable pace for six minutes between two points marked ten meters apart.^30^ The number of laps was recorded and as soon as the timer ended, participants stopped in their final location. A telescopic measuring device was utilized to obtain any additional distance between the end points and the participant’s final location.

The final walking assessment included three timed up-and-go (TUG) trials. Participants stood up from a chair, walked three meters forward, turned around, and returned back to a sitting position. The time taken to complete the trial was measured with a hand-held stopwatch.^31^ Participants were instructed to limit using their hands on the chair for support. Participants could use assistive walking devices such as a cane or walker, but wheelchairs were excluded. Participants were instructed to prioritize their safety and encouraged to stop and take breaks if necessary.

The Berg Balance Scale single leg test (*i.e.,* standing on one leg, BBS-SLT) was used clinically to assess balance and potential fall risk.^32, 33^ Participants were instructed to balance on one leg (paretic leg for stroke participants, matched-paretic leg for controls) for as long as possible, repeating the task three times on each leg. An experimenter stood with the participant and monitored their movements to prevent injuries. One stroke survivor chose not to participate in this assessment, while two others opted not to test their affected side. The balance scale for paretic limb was scored with 5 levels (scored from 0 to 4 points) and then dichotomized into scores of 0, 1, and 2 considered poor balance function, and scores of 3 and 4 considered to have acceptable or good balance.

### Data Management and Statistical Analyses

Statistical analysis was performed using IBM Statistical Product and Service Solutions (SPSS, Version 29). The following upper limb neuromotor capacities were computed: grip strength of both limbs (Newtons), grip strength symmetry (paretic side ÷ nonparetic side X 100), mean grip strength between limbs, simple reaction time of both limbs (ms), SRT symmetry, mean SRT between limbs, All Accuracy (correct go/no-go trials ÷ total go/no-go trials X100), All Accuracy symmetry, mean All Accuracy between limbs, All Accuracy/SRT of both limbs, All Accuracy/SRT symmetry, and mean All Accuracy/SRT between limbs. All Accuracy/SRT metrics were included as it rewards accuracy of decision-making and processing speed. For age-matched control participants, paretic and nonparetic limbs were treated as the matched-paretic and matched-nonparetic limbs, respectively. The continuous data were evaluated for normality.

Within participant and group comparisons for the non-normalized neuromotor capacities (*i.e.,* raw values) were evaluated with a 2X2 mixed measures analysis of variance (ANOVA) (Figure 2). Between group comparisons for the normalized neuromotor capacities (*e.g.,* symmetry, mean between limbs) and mobility and balance measures were evaluated with a multivariate ANOVA (MANOVA). The participants’ group (Stroke, Control) was used as the between-subjects factor, and the limb (paretic/matched-paretic, nonparetic/matched-nonparetic) was used as the within-subjects factor in the 2X2 ANOVA. A significant main or interaction effect was followed by a post-hoc comparison using independent or paired sample *t*-tests with Šidák corrections for multiple comparisons. For the MANOVA, the participants’ group was used as the between-subjects factor and the following variables were treated as the dependent factors: symmetry and mean scores for grip strength, SRT, All Accuracy, and All Accuracy/SRT as well as the mobility assessments (gait speed, 6MWT, TUG, BBS-SLT).

**Figure 2:**
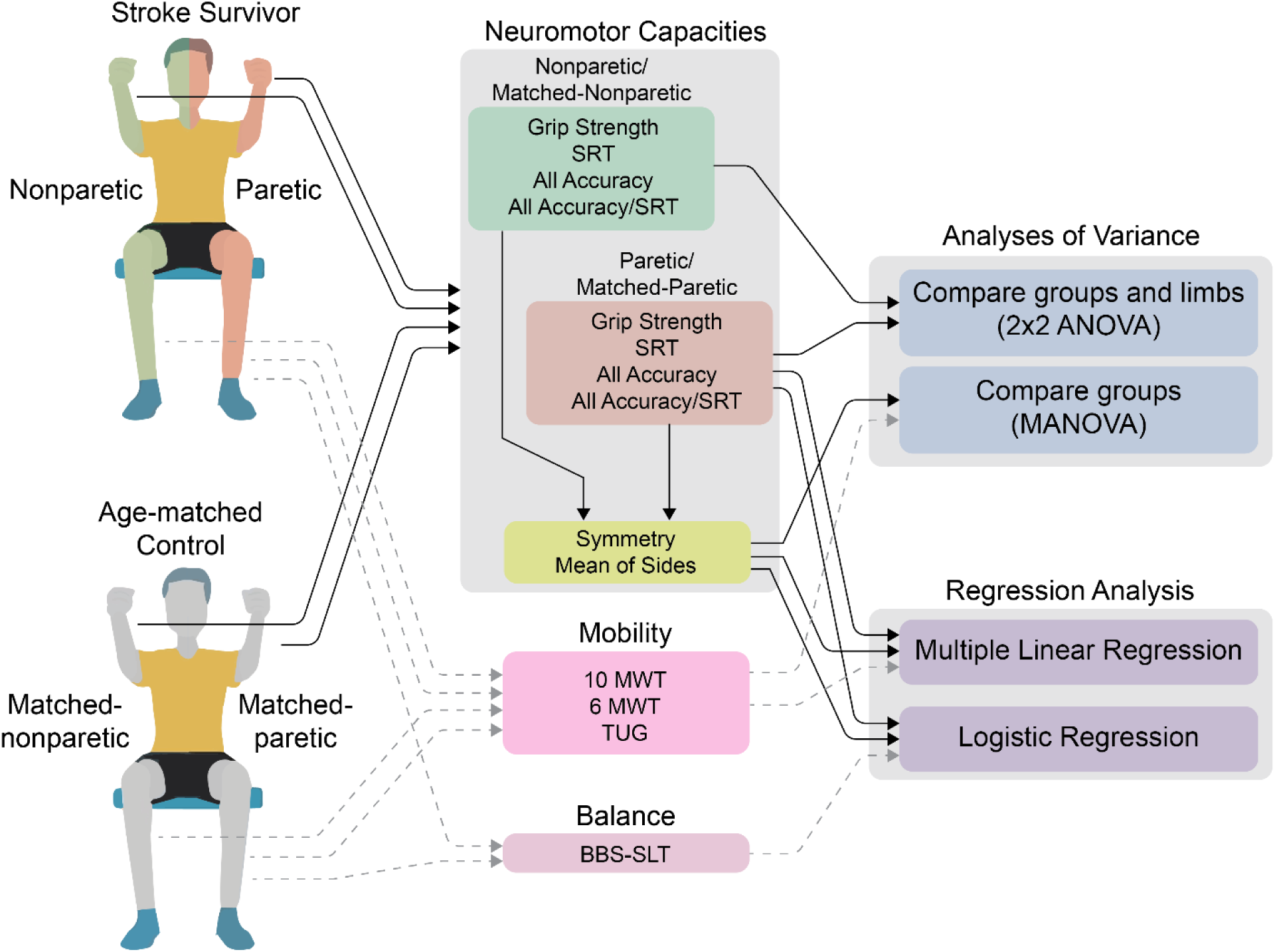
Measurements of neuromotor capacities (grip strength, simple reaction time [SRT], accuracy of both go and no-go reaction [All Accuracy], and All Accuracy/SRT) were collected from both upper extremities of each participant. Mobility measures (10 meter walk test [10 MWT], 6 minute walk test [6 MWT], and timed up-and-go [TUG]) and single-leg balance (BBS-SLT) were also collected. Normalized neuromotor capacity were computed using the raw capacities from both limbs. Between-limb and between-group comparisons were made using the raw neuromotor capacities, while only between-group comparisons were made using the normalized neuromotor capacities. Multiple linear regression was used to evaluate the normalized neuromotor capacities as predictors of the mobility measures, and logistic regression was used to evaluate the normalized neuromotor capacities as predictors of balance (Poor/Good)

We also performed a stepwise multiple linear regression analysis to evaluate how the computed neuromotor capacities that involved the paretic side (paretic limb scores, symmetry scores, and mean scores for grip strength, SRT, All Accuracy, and All Accuracy/SRT) predicted gait speed (m/s), timed up and go time (s), and six minute walk test distance (m). For BBS-SLT, the same neuromotor capacities were entered into a binary logistic regression model. Participants’ age was also included as a variable in the regression analysis. We also evaluated the relationships between the independent and continuous dependent variables with Pearson’s correlation coefficients (see Supplemental Material). All analyses were performed for the entire cohort and, separately, for the stroke group. A significance level of a=0.05 was used for all analyses.

## Results

### Descriptive Statistics

All independent and dependent variables were normally distributed except for TUG which was skewed due to a single outlier. Analyses were therefore performed with and without this data point and there was no substantive change in results. Reported results for TUG include all data points. The means and standard deviations for the neuromotor capacity metrics and mobility and balance measures are provided in Table 2 and 3. The correlations between these measures can be found in the Supplemental Material.

**Table 2:** Neuromotor Capacities.

| Neuromotor Capacity | Limb/ Normalization | Stroke | Control |
| --- | --- | --- | --- |
| Grip Strength (N) | P/M-P | 196.34 ± 95.74 | 356.97 ± 109.21 |
|  | NP/M-NP | 272.35 ± 99.29 | 351.9 ± 113.33 |
|  | Symmetry | 72.52 ± 25.07 | 101.91 ± 5.73 |
|  | Mean | 234.34 ± 89.71 | 354.44 ± 110.65 |
| Simple Reaction Time (ms) | P/M-P | 236.24 ± 42.78 | 200.89 ± 20.93 |
|  | NP/M-NP | 213.61 ± 26.05 | 206.01 ± 27.51 |
|  | Symmetry | 110.46 ± 13.6 | 98.15 ± 8.22 |
|  | Mean | 224.92 ± 32.26 | 203.45 ± 22.67 |
| All Accuracy (% correct) | P/M-P | 0.7 ± 0.14 | 0.77 ± 0.14 |
|  | NP/M-NP | 0.74 ± 0.11 | 0.78 ± 0.1 |
|  | Symmetry | 94.57 ± 15.36 | 99.12 ± 17.71 |
|  | Mean | 0.72 ± 0.11 | 0.77 ± 0.1 |
| All Accuracy/SRT (% correct/ms) | P/M-P | 0.30 ± 0.05 | 0.38 ± 0.07 |
|  | NP/M-NP | 0.35 ± 0.05 | 0.38 ± 0.06 |
|  | Symmetry | 86.16 ± 13.59 | 101.12 ± 17.79 |
|  | Mean | 0.32 ± 0.05 | 0.38 ± 0.06 |

Values are presented as mean ± standard deviation. P/M-P: Paretic/Matched-Paretic, NP/M-NP: Nonparetic/Matched-Nonparetic

**Table 3:** Mobility and Balance Measures.

| Mobility/Balance Measure | Stroke | Control |
| --- | --- | --- |
| 10 MWT (m/s) | 1.00 ± 0.36 | 1.27 ± 0.19 |
| TUG (s) | 13.68 ± 8.52 | 8.96 ± 1.72 |
| 6 MWT (min) | 273.75 ± 113.74 | 365.3 ± 50.06 |
| BBS-SLT<br>(Poor Balance/Total) | 14/20 | 1/20 |

10 MWT: 10 meter walk test, TUG: Timed Up-and-Go, 6 MWT: 6 minute walk test, BBS-SLT: Berg Balance Scale-Single Leg Test

### Neuromotor Capacities (Raw Values)

The 2X2 mixed measures ANOVAs indicated significant groupXlimb interaction for all variables (8.203 < *F_1,38_* <20.440, all *p*≤0.007) except for All Accuracy (*p*=0.461). There was also no significant main effects of group or limb for All Accuracy (*p*=0.111 and 0.180, respectively).

Interaction: Between Limb Comparisons within Each Group (Table 2):

Post-hoc analyses of groupXlimb interaction comparing limbs within each group revealed that the measures between limbs differed in Stroke but not in the Control group. Specifically, grip strength was reduced in the paretic limb as compared to the nonparetic limb (Δgrip strength = 76.0±12.7N, *p*<0.001), SRT was increased in the paretic limb as compared to the nonparetic limb (ΔSRT = 22.6±5.4ms, *p*<0.001), and All Accuracy/SRT was reduced in the paretic limb as compared to the nonparetic limb (ΔAll Accuracy/SRT = 0.05±0.01 % correct/ms, *p*<0.001). No differences between limbs were present in the Control group (all *p* > 0.354).

Interaction: Between Group Comparisons within Each Limb (Table 2 and 3):

Post-hoc analyses of groupXlimb interaction comparing groups within each limb revealed that all measures except for All Accuracy differed between groups in the paretic/matched-paretic limb, but only grip strength differed in the nonparetic/matched-nonparetic limb. Specifically, in the paretic/matched-paretic limb, grip strength was reduced in the Stroke group (Δgrip strength=160.6±32.5N, *p*<0.001), SRT was increased in the paretic limb (ΔSRT = 35.3±10.7ms, *p*=0.002), and All Accuracy/SRT was reduced (ΔAll Accuracy/SRT=0.09±0.02 %correct/ms, *p*<0.001). In the nonparetic/matched-nonparetic limb, grip strength was also reduced in the Stroke group (Δgrip strength=79.6.6+33.7N, *p*=0.023).

### Neuromotor Capacities (normalized) and Mobility and Balance Measures

The MANOVA indicated significant multivariate main effect of group (*F_12,27_=*4.637*, p=*0.001). The tests of between-subjects effects indicated that all variables were significantly impaired in the Stroke group when compared to the age- and sex-matched Control group (5.897 < *F_1,38_* < 31.175, all *p*<0.020) except for All Accuracy Symmetry (*F_1,38_=*0.756, *p*=0.390) and All Accuracy Mean of Sides (*F_1,38_=*2.667, *p*=0.111) (Table 2).

### Predictors of Mobility and Balance Measures

In the stroke group, stepwise multiple linear regression analysis revealed that grip strength symmetry was the only significant predictor of gait speed (R^2^=0.325, standardized slope *β_STD_* =0.570, *p*=0.009), timed up-and-go (R^2^=0.349, *β* =-0.591, *p*=0.006), and 6MWT (R^2^=0.290, *β_STD_* =0.539, *p*=0.014). Similarly, in the entire cohort, grip strength symmetry was a significant predictor of timed up-and-go (R^2^=0.405, *β_STD_* =-0.637, *p*<0.001) and 6MWT (R^2^=0.389, *β_STD_* =0.624, *p*<0.001). Further, grip strength symmetry, mean SRT, and All Accuracy symmetry were significant predictors of gait speed in the entire cohort (R^2^=0.524, Grip Strength symmetry *β_STD_* =0.429, mean SRT *β_STD_* =-0.377, All Accuracy symmetry *β_STD_* =0.245, *p*<0.001).

For balance, 6 out of 20 stroke survivors and 19 out of 20 age-matched controls were categorized as having good balance. In the stroke group, binary logistic regression analysis revealed that SRT symmetry was the only significant predictor of balance (Nagelkerke R^2^ = 0.530, exp(*β*) = 0.844, *p* = 0.002). Specifically, SRT symmetry was able to distinguish stroke survivors with good and poor balance with an odds ratio of 0.844, indicating that for each unit of increase in SRT symmetry (*i.e.,* longer SRT in the paretic limb), the odds of having good balance decreases by ≈16%. In the entire cohort, SRT symmetry and paretic/matched-paretic All Accuracy/SRT were the only significant predictors of balance (Nagelkerke R^2^ = 0.770, SRT symmetry exp(*β*) = 0.850, All Accuracy/SRT exp(*β*) = 48.701, *p* = 0.002). Specifically, SRT symmetry was able to distinguish all participants with good and poor balance with an odds ratio of 0.850, indicating that for each unit of increase in SRT symmetry, the odds of having good balance decreases by ≈15%. Additionally, paretic/matched-paretic All Accuracy/SRT was able to distinguish all participants with good and poor balance with an odds ratio of 48.701, indicating that for each 0.1 unit (*i.e.,* % correct/ms) increase in All Accuracy/SRT, the odds of having good balance increases by ≈477% (note that one unit increase in All Accuracy/SRT is not feasible).

## Discussion

This study demonstrated that the evaluation of upper limb neuromotor capacities including grip strength, reaction time (SRT), and short latency decision-making (All Accuracy and All Accuracy/SRT) in stroke survivors meeting study inclusion criteria is feasible, even for the paretic limb. Within-subject inter-limb comparisons demonstrated that the paretic limb had diminished grip strength, prolonged reaction time, and reduced speed–accuracy ratio (All Accuracy/SRT) in stroke survivors but not in controls. Between group differences showed that stroke survivors exhibited a reduction in grip strength in both limbs and slower reaction time and reduced speed–accuracy ratio in the paretic limb as compared with the matched limbs in the controls. Additionally, all normalized neuromotor capacities and mobility and balance measures were significantly impaired in the stroke survivors when compared with the controls except for All Accuracy Symmetry and All Accuracy Mean of Sides. There were significant correlations between many of the upper limb neuromotor capacities and the three measures of mobility (gait speed, TUG, and 6 MWT). Perhaps of greater interest, regression analyses demonstrated that grip strength symmetry was the primary independent predictor of these outcomes in both stroke survivors and the entire cohort. Additionally, the mean SRT and All Accuracy symmetry predicted gait speed in the entire cohort. In contrast, balance (BBS-SLT) was primarily predicted by SRT symmetry (in both stroke and the entire cohort) and paretic All Accuracy/SRT (only in the entire cohort). Age did not contribute to measures of mobility or balance in the presence of upper limb neuromotor capacities, underscoring the potency of these relationships. These findings highlight the utility of simple measures of neuromotor capacity like grip strength and reaction time in assessing post-stroke balance and mobility. These measures could serve as a valuable tool to inform clinical decision-making in stroke rehabilitation.

Is there a rationale for upper limb neuromotor strength, speed, and accuracy relating to accepted clinical measures of mobility and balance? It is not likely that distal upper limb function is directly responsible for this relationship given the near negligible role of the wrist and hand in maintaining gait and balance, as well as the lack of reported effects on gait in stroke survivors who regularly receive botulinum toxin in the upper extremity. Therefore, it is more likely that the relationship between upper limb function and gait/balance primarily driven by central factors. For instance, we observed that the extent to which grip strength was reduced in the paretic limb (grip strength symmetry) predicted all mobility measures. Previous studies have shown that upper extremity impairment following a stroke is driven by the integrity of the corticospinal tract (CST) by demonstrating that the presence of motor evoked potentials in the more-impaired limb, triggered by stimulation of the ipsilesional motor cortex, predict clinical measures of impairment including muscle strength.^34, 35^ However, the CST also contributes to motor control in the trunk and lower extremities.^36^ Following this, grip strength symmetry could be considered as a general indicator of post-stroke impairment rather than only as a measure of distal upper extremity strength. This interpretation more closely matches how grip strength is interpreted in other fields of rehabilitation, such as gerontology, where declines in grip strength with aging are less due to loss of peripheral muscle and more related to central changes leading to desynchronization of the central networks that also affect gait speed.^5, 37–39^ What is novel to this research is that grip strength symmetry, rather than simple grip strength, was the strongest independent predictor of mobility measures in the group as a whole and in the stroke survivors. Given what is known about grip strength, grip symmetry is likely an effective surrogate for the degree to which the stroke event impacted networks essential to walking within the injured hemisphere.

Interestingly, SRT was the only significant predictor of balance in stroke survivors, while the neuromotor capacities involving grip strength did not improve balance prediction. The speed– accuracy ratio (All Accuracy/SRT) was also a significant predictor of balance in the entire cohort; however, this was likely driven by the contribution of reaction time to this measure because All Accuracy was minimally affected by stroke and did not differ between groups. This finding aligns with previous studies that have examined the relationship between SRT and fall risk in individuals acute stroke.^40^ Based on our findings, it appears that inhibitory processes associated with correct response selection is minimally affected by stroke, and therefore, the primary contribution to poorer balance after stroke is slower activation of a selected motor plan rather than the inhibitory process associated with the correct selection of the plan.

Is there a mechanistic explanation for why stroke would affect reaction time but not accuracy? SRT relies on frontal cortical resources for attention as well as neural pathways associated with premotor cortical excitation of the basal ganglia, thalamocortical drive, and the corticospinal tract.^41^ Prior work has found that the speed–accuracy ratio is a potent predictor of response to sagittal and frontal perturbations during gait.^19, 22, 24^ Here, the ability to quickly inhibit an action, as evaluated through reaction accuracy, is an advantage. For example, modifying gait in response to a perturbation first requires inhibition of the pre-potent, or pre-planned, step which becomes obsolete at the instant of perturbation. This inhibition is thought to be a prerequisite for initiating a more adaptive stepping pattern.^42^ There is evidence that short latency inhibition is dependent on a direct white matter tract from frontal cortical regions to the sub-thalamic nucleus which then stimulates the globus pallidus to release GABA, an inhibitory neurotransmitter, to the basal ganglia which then attenuates thalamo-cortical drive and the pre-planned movement.^43^ This must occur quickly to allow successful response to perturbation and so likely relies on white matter integrity. However, our findings suggest that stroke does not significantly impair this inhibitory mechanism, pointing instead to slower motor activation as the primary issue affecting balance post-stroke. Given that SRT is driven in part by corticospinal activation and our finding that grip strength symmetry, also a measure of corticospinal tract integrity, predicted mobility measures, it is likely that both observations are the result of damage to the corticospinal tract resulting from the stroke. However, it is important to note that the relative contributions of inhibition and motor activation to neuromotor capacities could relate to lesion location, which was not considered in this study.

### Clinical Implications

Our finding that upper extremity neuromotor capacities can predict mobility and balance have meaningful implications for stroke rehabilitation. Specifically, these findings suggest that targeting muscle weakness and reaction time during therapy would lead to improvements in gait function and balance. Although the value of strength training in rehabilitation is debated, as gains are not believed to transfer well to functional activities,^44, 45^ our findings show that reduced strength limits mobility, suggesting that improvements in strength would have meaningful effects on function. Specifically, functional resistance training—integrating resistance training into functional activities like walking—could potentially facilitate the transfer of strength gains to activities of daily living. Similarly, reaction time training may have value in improving balance and minimizing fall risk in stroke survivors. Although this is an understudied topic in stroke, reaction training is commonplace in athletic training to improve sports performance. Again, it is to be noted that such training should be performed in a functional manner to better translate reaction improvements to function.^46^ Furthermore, upper limb neuromotor capacities may provide prognostic value. For instance, grip strength symmetry could provide prognostic information regarding the likelihood of gait recovery in people with stroke prior to rehabilitation efforts, similar to how grip strength can be used predict response to gait rehabilitation in frail older adults.^47^ Similarly, SRT may predict successful response to perturbation and balance and might be useful in prognostication when developing realistic rehabilitation goals regarding balance and fall risk in the setting of stroke. However, future work is necessary to fully understand the prognostic value of neuromotor capacities in stroke.

## Conclusions

In summary, the results of this study indicate that upper limb neuromotor measures of muscle strength, reaction time, and reaction accuracy can feasibly be obtained in stroke survivors. We observed strong relationships between grip strength symmetry and all measures of mobility as well as between simple reaction time symmetry and balance. Additionally, although grip strength symmetry has been associated with some important mobility outcomes in older people without hemiparesis, this is the first study to evaluate this parameter in stroke survivors. Functional balance on the paretic limb in the stroke survivors was best predicted by SRT symmetry. Grip strength and reaction time measures are clinically feasible and offer minimal risk methods for evaluating gait and balance potential in stroke survivors. Additionally, muscle strength and reaction time should be addressed in clinic to improve gait and balance after stroke. With further work, these measures may have utility in prognostication of rehabilitation outcomes related to mobility and balance in stroke survivors. The results are also of theoretical interest in that they offer a window into the integrity of neural pathways.

## Data Availability

The data that support the findings of this study are available from the corresponding author upon reasonable request.

## Acknowledgements

Drs. Augenstein and Krishnan had full access to all data and are responsible for the integrity of the data and accuracy of the analysis

## Non-standard Abbreviations and Acronyms

RT: Reaction Time
SRT: Simple Reaction Time
10MWT: 10 meter walk test
6MWT: 6 minute walk test
TUG: timed up-and-go
MMSE: Mini-mental state examination
NI: National Instruments
BBS-SLT: Berg Balance Scale-Single Leg Test
CST: corticospinal tract

