## Supplemental Material for "Predicting Post-Stroke Gait and Balance Function with Simple Neuromotor Measures"

| Gait speed (stroke) | | | | | | | | | | | | | | | |
| --- | --- | --- | --- | --- | --- | --- | --- | --- | --- | --- | --- | --- | --- | --- | --- |
|  |  |  | Grip Strength | | | SRT | | | All Accuracy | | | All Accuracy/SRT | | |  |
|  |  | Gait speed | P/M-P | Symm | Mean | P/M-P | Symm | Mean | P/M-P | Symm | Mean | P/M-P | Symm | Mean | Age |
|  | Gait speed | 1.00 | 0.36 | 0.57 | 0.19 | -0.53 | -0.28 | -0.56 | -0.19 | -0.02 | -0.24 | 0.29 | 0.20 | 0.21 | 0.04 |
| Grip Strength | P/M-P |  | 1.00 | 0.66 | 0.92 | -0.58 | -0.40 | -0.57 | -0.14 | -0.14 | -0.11 | 0.37 | 0.17 | 0.34 | 0.22 |
|  | Symm |  |  | 1.00 | 0.32 | -0.63 | -0.55 | -0.57 | -0.13 | -0.27 | -0.03 | 0.46 | 0.15 | 0.43 | 0.22 |
|  | Mean |  |  |  | 1.00 | -0.44 | -0.18 | -0.48 | -0.11 | -0.04 | -0.13 | 0.26 | 0.11 | 0.26 | 0.14 |
| SRT | P/M-P |  |  |  |  | 1.00 | 0.73 | 0.96 | 0.55 | 0.54 | 0.42 | -0.33 | 0.00 | -0.39 | -0.26 |
|  | Symm |  |  |  |  |  | 1.00 | 0.52 | 0.31 | 0.40 | 0.19 | -0.36 | -0.34 | -0.21 | -0.17 |
|  | Mean |  |  |  |  |  |  | 1.00 | 0.58 | 0.52 | 0.47 | -0.25 | 0.14 | -0.38 | -0.24 |
| All Accuracy | P/M-P |  |  |  |  |  |  |  | 1.00 | 0.72 | 0.92 | 0.61 | 0.51 | 0.43 | 0.10 |
|  | Symm |  |  |  |  |  |  |  |  | 1.00 | 0.38 | 0.26 | 0.72 | -0.10 | -0.04 |
|  | Mean |  |  |  |  |  |  |  |  |  | 1.00 | 0.65 | 0.26 | 0.63 | 0.17 |
| All Accuracy/SRT | P/M-P |  |  |  |  |  |  |  |  |  |  | 1.00 | 0.56 | 0.87 | 0.34 |
|  | Symm |  |  |  |  |  |  |  |  |  |  |  | 1.00 | 0.08 | 0.26 |
|  | Mean |  |  |  |  |  |  |  |  |  |  |  |  | 1.00 | 0.33 |
|  | Age |  |  |  |  |  |  |  |  |  |  |  |  |  | 1.00 |

P/M-P: Paretic/Matched-Paretic, Symm: Symmetry, Mean: Mean of Sides

| Gait speed (entire cohort) | | | | | | | | | | | | | | | |
| --- | --- | --- | --- | --- | --- | --- | --- | --- | --- | --- | --- | --- | --- | --- | --- |
|  |  |  | Grip Strength | | | SRT | | | All Accuracy | | | All Accuracy/SRT | | |  |
|  |  | Gait speed | P/M-P | Symm | Mean | P/M-P | Symm | Mean | P/M-P | Symm | Mean | P/M-P | Symm | Mean | Age |
|  | Gait speed | 1.00 | 0.52 | 0.63 | 0.42 | -0.57 | -0.32 | -0.58 | 0.12 | 0.16 | 0.06 | 0.50 | 0.34 | 0.45 | -0.08 |
| Grip Strength | P/M-P |  | 1.00 | 0.65 | 0.97 | -0.59 | -0.46 | -0.53 | 0.28 | 0.21 | 0.24 | 0.66 | 0.50 | 0.57 | -0.03 |
|  | Symm |  |  | 1.00 | 0.44 | -0.68 | -0.63 | -0.56 | 0.08 | -0.06 | 0.15 | 0.52 | 0.33 | 0.49 | 0.08 |
|  | Mean |  |  |  | 1.00 | -0.50 | -0.33 | -0.48 | 0.29 | 0.26 | 0.22 | 0.60 | 0.47 | 0.52 | -0.06 |
| SRT | P/M-P |  |  |  |  | 1.00 | 0.67 | 0.94 | 0.21 | 0.23 | 0.15 | -0.50 | -0.20 | -0.54 | -0.11 |
|  | Symm |  |  |  |  |  | 1.00 | 0.39 | 0.13 | 0.25 | 0.01 | -0.35 | -0.38 | -0.23 | -0.10 |
|  | Mean |  |  |  |  |  |  | 1.00 | 0.21 | 0.17 | 0.19 | -0.47 | -0.09 | -0.56 | -0.06 |
| All Accuracy | P/M-P |  |  |  |  |  |  |  | 1.00 | 0.75 | 0.90 | 0.73 | 0.63 | 0.60 | -0.08 |
|  | Symm |  |  |  |  |  |  |  |  | 1.00 | 0.39 | 0.49 | 0.79 | 0.20 | -0.06 |
|  | Mean |  |  |  |  |  |  |  |  |  | 1.00 | 0.69 | 0.37 | 0.69 | -0.06 |
| All Accuracy/SRT | P/M-P |  |  |  |  |  |  |  |  |  |  | 1.00 | 0.69 | 0.92 | 0.02 |
|  | Symm |  |  |  |  |  |  |  |  |  |  |  | 1.00 | 0.34 | 0.06 |
|  | Mean |  |  |  |  |  |  |  |  |  |  |  |  | 1.00 | 0.00 |
|  | Age |  |  |  |  |  |  |  |  |  |  |  |  |  | 1.00 |

P/M-P: Paretic/Matched-Paretic, Symm: Symmetry, Mean: Mean of Sides

| TUG (stroke) | | | | | | | | | | | | | | | |
| --- | --- | --- | --- | --- | --- | --- | --- | --- | --- | --- | --- | --- | --- | --- | --- |
|  |  |  | Grip Strength | | | SRT | | | All Accuracy | | | All Accuracy/SRT | | |  |
|  |  | TUG | P/M-P | Symm | Mean | P/M-P | Symm | Mean | P/M-P | Symm | Mean | P/M-P | Symm | Mean | Age |
|  | TUG | 1.00 | -0.36 | -0.59 | -0.19 | 0.44 | 0.13 | 0.50 | 0.18 | 0.21 | 0.12 | -0.23 | 0.09 | -0.30 | 0.04 |
| Grip Strength | P/M-P |  | 1.00 | 0.66 | 0.92 | -0.58 | -0.40 | -0.57 | -0.14 | -0.14 | -0.11 | 0.37 | 0.17 | 0.34 | 0.22 |
|  | Symm |  |  | 1.00 | 0.32 | -0.63 | -0.55 | -0.57 | -0.13 | -0.27 | -0.03 | 0.46 | 0.15 | 0.43 | 0.22 |
|  | Mean |  |  |  | 1.00 | -0.44 | -0.18 | -0.48 | -0.11 | -0.04 | -0.13 | 0.26 | 0.11 | 0.26 | 0.14 |
| SRT | P/M-P |  |  |  |  | 1.00 | 0.73 | 0.96 | 0.55 | 0.54 | 0.42 | -0.33 | 0.00 | -0.39 | -0.26 |
|  | Symm |  |  |  |  |  | 1.00 | 0.52 | 0.31 | 0.40 | 0.19 | -0.36 | -0.34 | -0.21 | -0.17 |
|  | Mean |  |  |  |  |  |  | 1.00 | 0.58 | 0.52 | 0.47 | -0.25 | 0.14 | -0.38 | -0.24 |
| All Accuracy | P/M-P |  |  |  |  |  |  |  | 1.00 | 0.72 | 0.92 | 0.61 | 0.51 | 0.43 | 0.10 |
|  | Symm |  |  |  |  |  |  |  |  | 1.00 | 0.38 | 0.26 | 0.72 | -0.10 | -0.04 |
|  | Mean |  |  |  |  |  |  |  |  |  | 1.00 | 0.65 | 0.26 | 0.63 | 0.17 |
| All Accuracy/SRT | P/M-P |  |  |  |  |  |  |  |  |  |  | 1.00 | 0.56 | 0.87 | 0.34 |
|  | Symm |  |  |  |  |  |  |  |  |  |  |  | 1.00 | 0.08 | 0.26 |
|  | Mean |  |  |  |  |  |  |  |  |  |  |  |  | 1.00 | 0.33 |
|  | Age |  |  |  |  |  |  |  |  |  |  |  |  |  | 1.00 |

P/M-P: Paretic/Matched-Paretic, Symm: Symmetry, Mean: Mean of Sides

| TUG (entire cohort) | | | | | | | | | | | | | | | |
| --- | --- | --- | --- | --- | --- | --- | --- | --- | --- | --- | --- | --- | --- | --- | --- |
|  |  |  | Grip Strength | | | SRT | | | All Accuracy | | | All Accuracy/SRT | | |  |
|  |  | TUG | P/M-P | Symm | Mean | P/M-P | Symm | Mean | P/M-P | Symm | Mean | P/M-P | Symm | Mean | Age |
|  | TUG | 1.00 | -0.41 | -0.64 | -0.30 | 0.50 | 0.25 | 0.50 | -0.02 | 0.01 | -0.04 | -0.35 | -0.16 | -0.36 | 0.08 |
| Grip Strength | P/M-P |  | 1.00 | 0.65 | 0.97 | -0.59 | -0.46 | -0.53 | 0.28 | 0.21 | 0.24 | 0.66 | 0.50 | 0.57 | -0.03 |
|  | Symm |  |  | 1.00 | 0.44 | -0.68 | -0.63 | -0.56 | 0.08 | -0.06 | 0.15 | 0.52 | 0.33 | 0.49 | 0.08 |
|  | Mean |  |  |  | 1.00 | -0.50 | -0.33 | -0.48 | 0.29 | 0.26 | 0.22 | 0.60 | 0.47 | 0.52 | -0.06 |
| SRT | P/M-P |  |  |  |  | 1.00 | 0.67 | 0.94 | 0.21 | 0.23 | 0.15 | -0.50 | -0.20 | -0.54 | -0.11 |
|  | Symm |  |  |  |  |  | 1.00 | 0.39 | 0.13 | 0.25 | 0.01 | -0.35 | -0.38 | -0.23 | -0.10 |
|  | Mean |  |  |  |  |  |  | 1.00 | 0.21 | 0.17 | 0.19 | -0.47 | -0.09 | -0.56 | -0.06 |
| All Accuracy | P/M-P |  |  |  |  |  |  |  | 1.00 | 0.75 | 0.90 | 0.73 | 0.63 | 0.60 | -0.08 |
|  | Symm |  |  |  |  |  |  |  |  | 1.00 | 0.39 | 0.49 | 0.79 | 0.20 | -0.06 |
|  | Mean |  |  |  |  |  |  |  |  |  | 1.00 | 0.69 | 0.37 | 0.69 | -0.06 |
| All Accuracy/SRT | P/M-P |  |  |  |  |  |  |  |  |  |  | 1.00 | 0.69 | 0.92 | 0.02 |
|  | Symm |  |  |  |  |  |  |  |  |  |  |  | 1.00 | 0.34 | 0.06 |
|  | Mean |  |  |  |  |  |  |  |  |  |  |  |  | 1.00 | 0.00 |
|  | Age |  |  |  |  |  |  |  |  |  |  |  |  |  | 1.00 |

P/M-P: Paretic/Matched-Paretic, Symm: Symmetry, Mean: Mean of Sides

| 6 MWT (stroke) | | | | | | | | | | | | | | | |
| --- | --- | --- | --- | --- | --- | --- | --- | --- | --- | --- | --- | --- | --- | --- | --- |
|  |  |  | Grip Strength | | | SRT | | | All Accuracy | | | All Accuracy/SRT | | |  |
|  |  | 6MWT | P/M-P | Symm | Mean | P/M-P | Symm | Mean | P/M-P | Symm | Mean | P/M-P | Symm | Mean | Age |
|  | 6MWT | 1.00 | 0.30 | 0.54 | 0.12 | -0.50 | -0.29 | -0.53 | -0.30 | -0.07 | -0.37 | 0.13 | 0.14 | 0.04 | -0.11 |
| Grip Strength | P/M-P |  | 1.00 | 0.66 | 0.92 | -0.58 | -0.40 | -0.57 | -0.14 | -0.14 | -0.11 | 0.37 | 0.17 | 0.34 | 0.22 |
|  | Symm |  |  | 1.00 | 0.32 | -0.63 | -0.55 | -0.57 | -0.13 | -0.27 | -0.03 | 0.46 | 0.15 | 0.43 | 0.22 |
|  | Mean |  |  |  | 1.00 | -0.44 | -0.18 | -0.48 | -0.11 | -0.04 | -0.13 | 0.26 | 0.11 | 0.26 | 0.14 |
| SRT | P/M-P |  |  |  |  | 1.00 | 0.73 | 0.96 | 0.55 | 0.54 | 0.42 | -0.33 | 0.00 | -0.39 | -0.26 |
|  | Symm |  |  |  |  |  | 1.00 | 0.52 | 0.31 | 0.40 | 0.19 | -0.36 | -0.34 | -0.21 | -0.17 |
|  | Mean |  |  |  |  |  |  | 1.00 | 0.58 | 0.52 | 0.47 | -0.25 | 0.14 | -0.38 | -0.24 |
| All Accuracy | P/M-P |  |  |  |  |  |  |  | 1.00 | 0.72 | 0.92 | 0.61 | 0.51 | 0.43 | 0.10 |
|  | Symm |  |  |  |  |  |  |  |  | 1.00 | 0.38 | 0.26 | 0.72 | -0.10 | -0.04 |
|  | Mean |  |  |  |  |  |  |  |  |  | 1.00 | 0.65 | 0.26 | 0.63 | 0.17 |
| All Accuracy/SRT | P/M-P |  |  |  |  |  |  |  |  |  |  | 1.00 | 0.56 | 0.87 | 0.34 |
|  | Symm |  |  |  |  |  |  |  |  |  |  |  | 1.00 | 0.08 | 0.26 |
|  | Mean |  |  |  |  |  |  |  |  |  |  |  |  | 1.00 | 0.33 |
|  | Age |  |  |  |  |  |  |  |  |  |  |  |  |  | 1.00 |

P/M-P: Paretic/Matched-Paretic, Symm: Symmetry, Mean: Mean of Sides

| 6 MWT (entire cohort) | | | | | | | | | | | | | | | |
| --- | --- | --- | --- | --- | --- | --- | --- | --- | --- | --- | --- | --- | --- | --- | --- |
|  |  |  | Grip Strength | | | SRT | | | All Accuracy | | | All Accuracy/SRT | | |  |
|  |  | 6MWT | P/M-P | Symm | Mean | P/M-P | Symm | Mean | P/M-P | Symm | Mean | P/M-P | Symm | Mean | Age |
|  | 6MWT | 1.00 | 0.47 | 0.62 | 0.35 | -0.56 | -0.39 | -0.53 | 0.04 | 0.14 | -0.03 | 0.43 | 0.37 | 0.33 | -0.12 |
| Grip Strength | P/M-P |  | 1.00 | 0.65 | 0.97 | -0.59 | -0.46 | -0.53 | 0.28 | 0.21 | 0.24 | 0.66 | 0.50 | 0.57 | -0.03 |
|  | Symm |  |  | 1.00 | 0.44 | -0.68 | -0.63 | -0.56 | 0.08 | -0.06 | 0.15 | 0.52 | 0.33 | 0.49 | 0.08 |
|  | Mean |  |  |  | 1.00 | -0.50 | -0.33 | -0.48 | 0.29 | 0.26 | 0.22 | 0.60 | 0.47 | 0.52 | -0.06 |
| SRT | P/M-P |  |  |  |  | 1.00 | 0.67 | 0.94 | 0.21 | 0.23 | 0.15 | -0.50 | -0.20 | -0.54 | -0.11 |
|  | Symm |  |  |  |  |  | 1.00 | 0.39 | 0.13 | 0.25 | 0.01 | -0.35 | -0.38 | -0.23 | -0.10 |
|  | Mean |  |  |  |  |  |  | 1.00 | 0.21 | 0.17 | 0.19 | -0.47 | -0.09 | -0.56 | -0.06 |
| All Accuracy | P/M-P |  |  |  |  |  |  |  | 1.00 | 0.75 | 0.90 | 0.73 | 0.63 | 0.60 | -0.08 |
|  | Symm |  |  |  |  |  |  |  |  | 1.00 | 0.39 | 0.49 | 0.79 | 0.20 | -0.06 |
|  | Mean |  |  |  |  |  |  |  |  |  | 1.00 | 0.69 | 0.37 | 0.69 | -0.06 |
| All Accuracy/SRT | P/M-P |  |  |  |  |  |  |  |  |  |  | 1.00 | 0.69 | 0.92 | 0.02 |
|  | Symm |  |  |  |  |  |  |  |  |  |  |  | 1.00 | 0.34 | 0.06 |
|  | Mean |  |  |  |  |  |  |  |  |  |  |  |  | 1.00 | 0.00 |
|  | Age |  |  |  |  |  |  |  |  |  |  |  |  |  | 1.00 |

P/M-P: Paretic/Matched-Paretic, Symm: Symmetry, Mean: Mean of Sides
